# Patterns of primary oral and maxillofacial malignancies among patients seen at Tikur Anbessa comprehensive specialized hospital – A retrospective study

**DOI:** 10.1101/2022.07.12.22277543

**Authors:** Chula Ararsa, Demerit Dejene, Gelana Garoma

**Author notes:** Corresponding author: (GG). These authors contributed equally to this work.

## Abstract

**Background:** Orofacial cancer is a malignant neoplastic proliferation of epithelial and ectomesenchymal tissue of oral and maxillofacial origin. The late presentation of patient, aggressive nature of orofacial malignancy and the anatomic site closure to vital organ make orofacial cancer management challenging.

**Objective:** The goal of this was to assess the patterns and risk factors of primary orofacial malignancy among patients visited Tikur Anbessa comprehensive specialized hospital.

**Patients and methods:** A cross sectional descriptive study with retrospective data collection was conducted on 175 patients diagnosed with primary oral and maxillofacial malignancies at Tikur Anbessa comprehensive specialized hospital over a period of January 2020 to December 2021. Data were collected by chart review. The collected date were entered to SPSS 25.0 for statistical analysis and results were presented with table, figures and charts. Percentage and frequency were employed for categorical data while mean was used for continuous variables.

**Results:** Out of 175 primary orofacial cases analysed, male were 57.1 %(n=100) with male to female ratio 1.48: 1, mean age of (48.21 ± 16.93 years) and range (12–91 years). Squamous cell carcinoma was the commonest cancer (52.0%) followed by mucoepidermoid carcinoma. About 34 % of patient had known risk factor. Majority of the patient (65.7%) were diagnosed as stage IV. Distance metastases were identified in 8.6 % of the patients and 41.7% of patients were treated surgically.

**Conclusion:** The study showed squamous cell carcinoma was the most prevalent orofacial cancer. Majority of patients were presented with advanced stage of disease. Surgery was the main means of treatment modality given to orofacial cancer patients.

## Introduction

Oral and maxillofacial cancer is a malignant neoplasia arising from structure confined in the oral and maxillofacial regions. Oral cancer is among the most common malignancies worldwide. Often it involves the oral mucosa and underlying structures in the area of oral cavity, lip, maxilla, mandible, facial skeleton, salivary gland, face and facial skin (1, 2). It constitute varying proportions of the total incidence of malignancies in the human population (3). The incidence of oral cancer and maxillofacial cancer is rising due to overindulgence in tobacco chewing and smoking. Oral cancer is identified as the 11^th^ most common malignancy in the world (4).

The aetiology of oral and maxillofacial malignancy is multifactorial and the major known risk factors are tobacco smoking and alcohol consumption (5, 6). Both of these factors account for nearly 90% of the cases and are associated with age, sex, and religion-ethnicity distribution (7). A number of other risk factor have been identified including human papilloma virus (HPPV 16 and 18), ionization radiation, chronic irritation, nutritional deficiency, premalignant lesions and condition, poor oral hygiene, low socio economic status, immune suppression (8,9). Habits, such as betel nut are other factors that were identified (10).

Orofacial cancer exhibits a multitude of growth and degree of aggressiveness which is primarily determined by histologic grade (11). It has various clinical presentations, including pain, soft tissue and bony swellings with ulcerations (12). Anatomically, oral cavity cancer classified into sub sites according to anatomic area involved. These anatomic sub sites include: oral tongue, palate, mucosal lip, retro molar trigone, floor of mouth and alveolar gingiva (13).

Oral and maxillofacial malignancies (OMM) is associated with significant cosmetic and functional limitations (15-16). In addition, both the presence of OMM and its treatment often result in significant deterioration in the patient’s quality of life (17). The standard of treatment for orofacial cancer is surgery as the first line treatment option. This surgical resection with or without postoperative adjuvant therapy. Combination postoperative radiation and chemo radiation therapy have resulted in improved survival. For advanced orofacial cancer cases, multimodality therapy is indicated (18).

Even though, Orofacial cancer incidence and distribution is well known in developed country the exact extent and incidence is not known in Africa as a general and in Ethiopia as particular. So this paper is aimed to give clue on the distribution of primary orofacial cancer seen at Tikur Anbessa comprehensive specialized hospital.

## Materials and methods

The study was conducted at Tikur Anbessa comprehensive specialized hospital, oral and maxillofacial surgery and oncology department, Addis Ababa, Ethiopia. Tikur Anbessa comprehensive specialized hospital (TACSH) is the government owned pioneer tertiary hospital equipped with 600 bed and serve as the only well-equipped oncologic centre of the country. It is a under Addis Ababa university college of health science which is a home to many specialty and sub specialty centre. Oral and maxillofacial surgery is one of recently started postgraduate department in AAU, college of health sciences.

Retrospective cross sectional study was employed on chart of patient with primary orofacial malignant tumour who has visited TACSH from January 1/ 2020 to December 30, 2021. Data was collected from patients’ medical records. For each patient, a number of variables were recorded including their demographic data, types and site of tumor, risk factors, duration of lesion, and treatment received. Charts with incomplete information was excluded. Data was collected, summarized, coded and entered to SPSS 25.0 computer program software. Frequency distribution tables, graph were used to represent the results.

## Ethical consideration

Permission of ethical issue and ethical clearance was obtained from research and ethics committee of department of Dentistry, College health science, AAU before the study. This research was conducted in full accordance with the World Medical Association Declaration of Helsinki. We confirmed that patients’ information remained confidential and data was anonymized and de-identified to analysis. Patient name was not recorded and patient information kept confidential.

## Results

### Socio demographic characteristic of study population

Out of 175 total patient with oral and maxillofacial cancer seen and analyzed, 100 were male (57.1 %) and 75 were female with male to female ratio of 1.48:1. Majority of the patients were in fifth decade age followed by fourth and seventh decade with mean age of 48.21 and SD =16.930, range 12-93 year (Table 1).

**Table 1.** Age and sex distribution of orofacial cancer patient seen at Tikur Anbessa comprehensive specialized hospital during January 2020 to December 2021, Addis Ababa, Ethiopia

| Variables | Category | Frequency (%) |
| --- | --- | --- |
| Sex | Female | 75(42.8%) |
|  | Male | 100(57.2%) |
| Age groups (years) | 11 – 20 | 5(2.85%) |
|  | 21 – 30 | 19(10.85%) |
|  | 31 – 40 | 34(19.42%) |
|  | 41 – 50 | 42(24%) |
|  | 51 – 60 | 34(19.42%) |
|  | 61 – 70 | 21(12%) |
|  | >70 | 20(11.42%) |
|  | Mean ± SD | 48.21 ± 16.93 |
|  | Range | 12 - 93 year |

### Histopathology diagnosis

According to histopathologic type squamous cell carcinoma was the most prevalent orofacial cancer 50.8% (n=89) followed by mucoepidermoid carcinoma (MECA) 14.2% (n =26), which was the commonest salivary gland cancer. Adenocystic carcinoma (ACC) was the second most prevalent salivary gland cancer 11(6.3%) (figure1).

**Figure 1.**
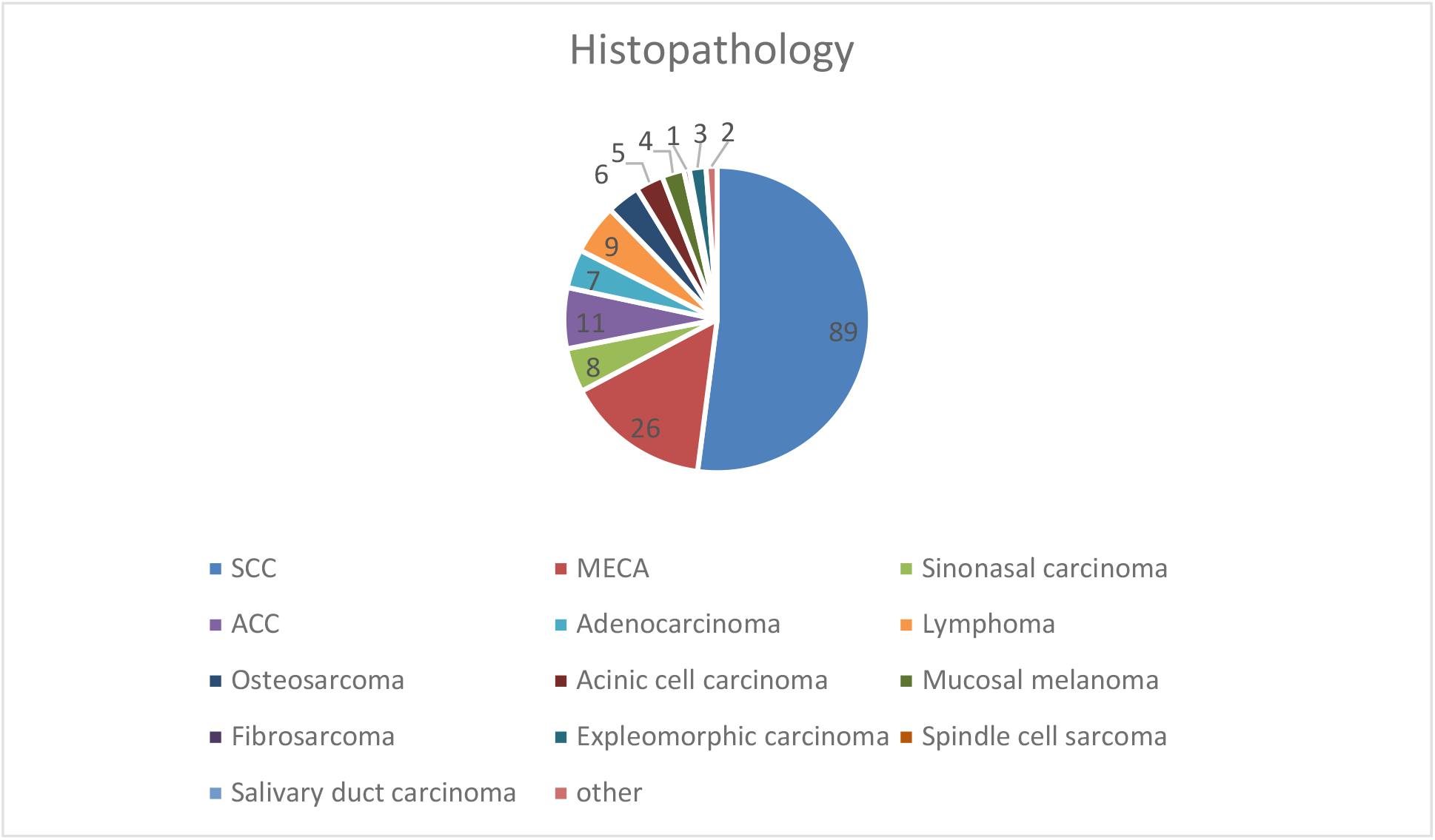
Histopathology distribution of orofacial cancer among patients seen at Tikur Anbessa comprehensive specialized hospital during January 2020 – December 2021, Addis Ababa, Ethiopia.

### Anatomic variation of Oral and maxillofacial cancer

Oral cavity were the most commonly affected anatomic site by orofacial cancer with 52.6% (n= 92) followed by parotid 14.3% and maxillary sinus 13.7%. Tongue was the commonly affected oral cavity sub-site with 20% (n=35) followed by lip and palate, 9.1 % (n=16) and 8.6% (n=15) respectively (Table 2).

**Table 2.** Anatomic distribution of orofacial cancer among patients seen at Tikur Anbessa comprehensive specialized hospital during January 2020-December 2021, Addis Ababa, Ethiopia

| Variables | Category | Frequency (%) |
| --- | --- | --- |
| Anatomic site | Tongue | 35(20%) |
|  | Maxillary sinus | 24 (13.7%) |
|  | Buccal mucosa | 14(8%) |
|  | Lip | 16(9.1%) |
|  | Parotid | 25(14.3%) |
|  | Palate | 15(8.6%) |
|  | Submandibular | 5(2.9%) |
|  | Retro molar | 3(1.7%) |
|  | Alveolar gingiva | 6(3.4%) |
|  | Maxilla | 12(6.9%) |
|  | Mandible | 8 (4.6%) |
|  | Floor of mouth | 3 (1.7%) |
|  | Cervical | 2 (1.1%) |
|  | Other | 7 (4%) |

### Stage of orofacial cancer

Majority of the patients were presented with advanced stage of disease with stage IV that accounted about 65.7%, stage III 11.7%. Almost half (49.1%) study subjects were with locally advanced tumor size of T4a. Out of studied patients 29.7% had N2 lymph nodes metastasis while 53.7% were negative neck, 8.6% of the patient had distance metastasis mainly to lung (Table 3).

**Table 3.** TNM stage distribution of orofacial cancer among patients seen at Tikur Anbessa comprehensive specialized hospital during January 2020-December 2021, Addis Ababa, Ethiopia.

| Variables | Category | Frequency (%) |
| --- | --- | --- |
| Staging | Stage I | 7(4%) |
|  | Stage II | 20(11.4%) |
|  | Stage III | 30(11.7%) |
|  | Stage IV | 115(65.7%) |
|  | Missing | 3(1.7%) |
| Tumor size | T1 | 4(2.3%) |
|  | T2 | 37(21.1%) |
|  | T3 | 33(18.9%) |
|  | T4a | 86(49.1%) |
|  | T4b | 8(4.6%) |
| Lymph node | N | 124(13.7%) |
|  | N2 | 52(29.7%) |
|  | N3 | 5(2.9%) |
|  | Negative | 94(53.7%) |
| Metastasis | M0 | 160(91.4%) |
|  | M | 15(8.6%) |

**Table 4.** Duration of presentation of orofacial cancer patients seen at Tikur Anbessa specialized hospital during January 2020 – December 2021, Addis Ababa, Ethiopia

| Variables | Category | Frequency (%) |
| --- | --- | --- |
| Duration | < 6 month | 31 (17.71%) |
|  | 6 month -1 year | 72 (41.14%) |
|  | 1 year-2 year | 30 (17.14%) |
|  | >2 years | 42 (24.0%) |
|  | Total | 175 (100%) |

### Duration of lesion

Majority of patients were presented on a mean time of between 6 month and 1year of lesion onset (41.1%) followed by lesion greater than 2 year (24.0%), < 6month (17.71%) and 1year −2 year (17.14%).

### Risk factor of orofacial cancer patient

Out of 175 case analyzed, only 32% of patient had known risk factor like smoking, immune compromising disease, alcohol and chat chewing. From this known risk factors immunocompromised individuals account of 13.7% followed by smoking 10.3% (Table 5).

**Table 5.**
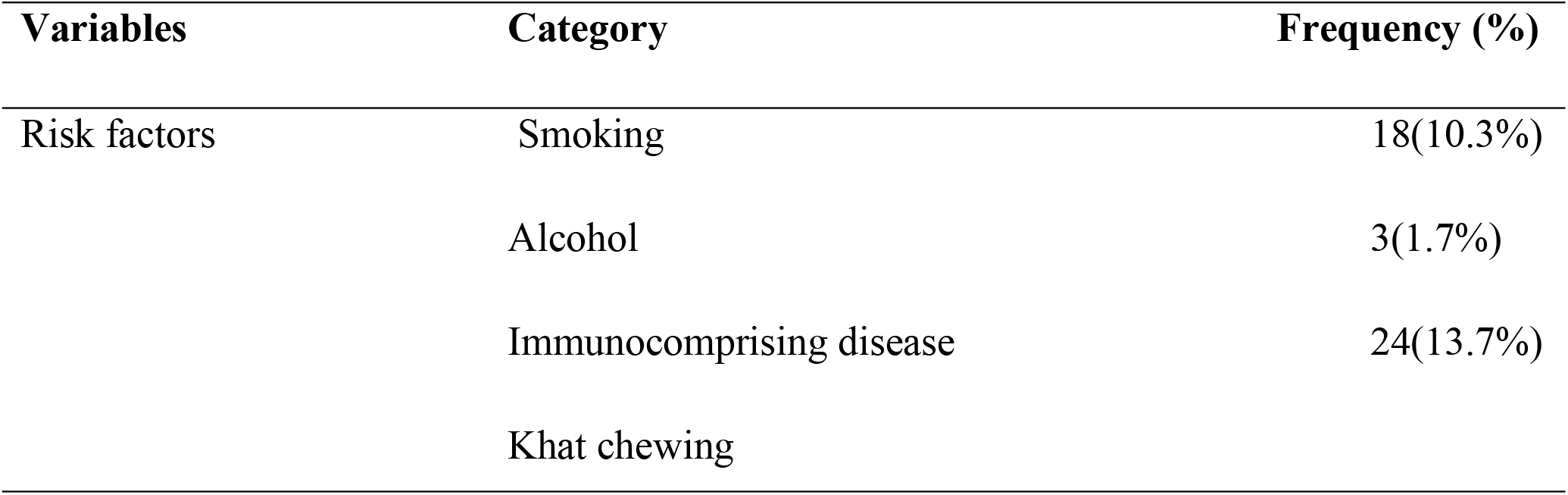
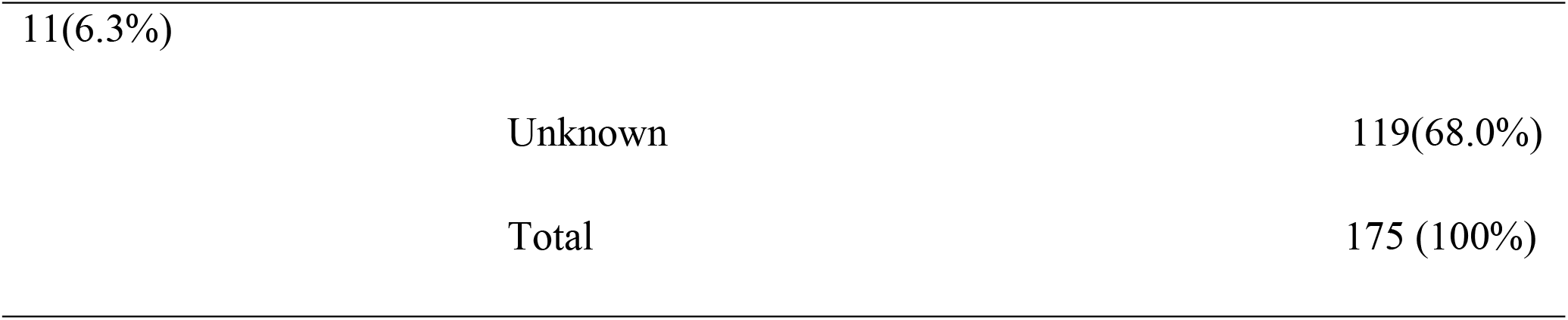
Risk factors and habits for orofacial cancer among patients seen at Tikur Anbessa comprehensive specialized hospital during January 2020 – December 2021, Addis Ababa, Ethiopia.

### Treatment modality

Surgery was the main mode of treatment given 73(41.7%), followed by chemotherapy 43(19.4%), surgery and chemo radiotherapy 5(2.9%), Radiotherapy 2(1.1%), while 61 (34.86%) of patient were not treated (Figure 2).

**Figure 2.**
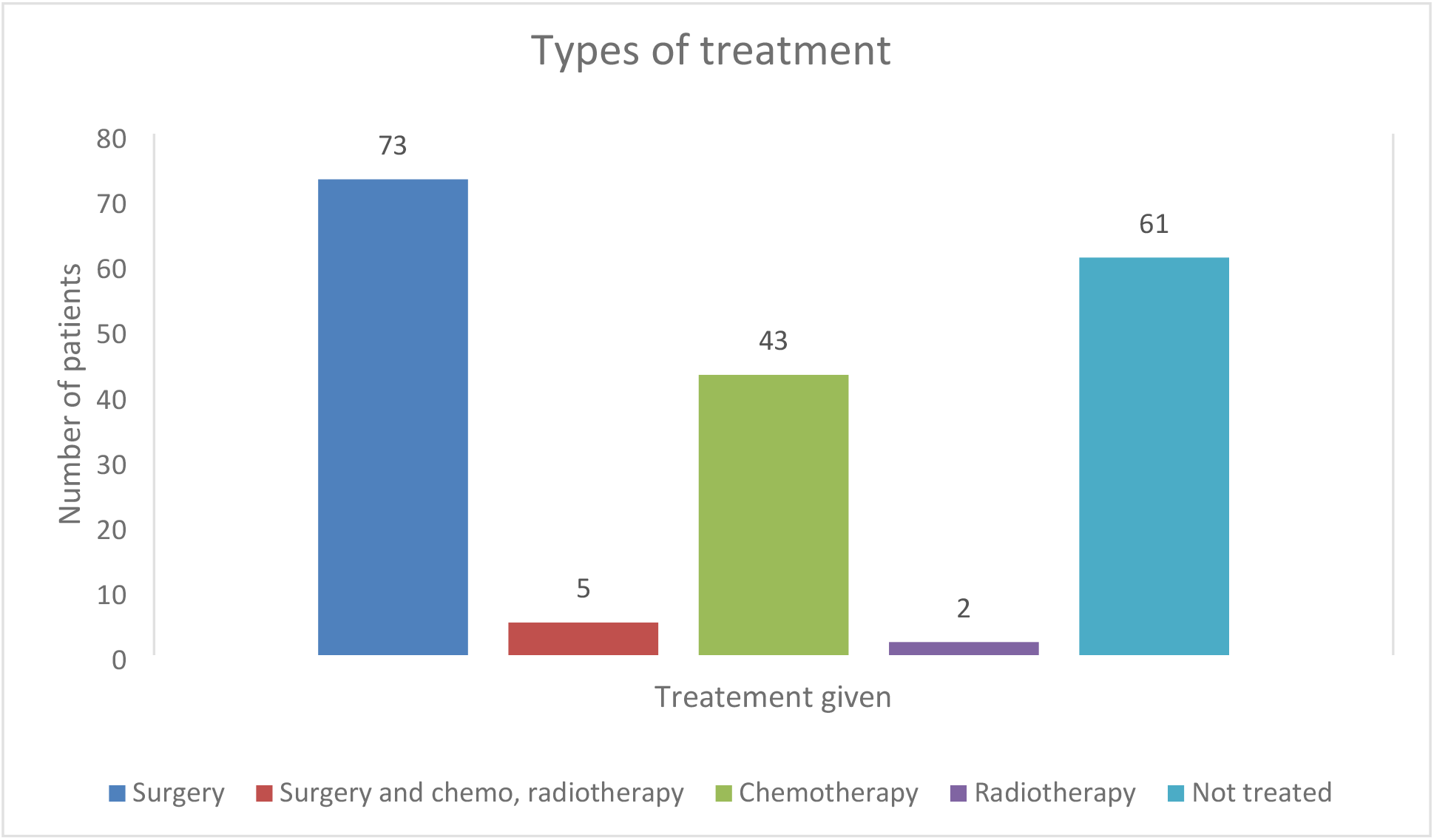
Type treatment of orofacial cancer among patients seen at Tikur Anbesa specialized Hospital during January 2020 – December 2021, Addis Ababa, Ethiopia.

## Discussion

Oral and maxillofacial cancer is a global disease seen in all age groups including children. In this study, out of total 175 patients the most common age group affected by oral and maxillofacial cancer were in fifth decade followed by fourth with mean age of 48.21±16.93 (range 12-93 years). The retrospective study done in Nigeria, by Adesina et al reported (48.7 ± 19.3 years) as the mean age and range (14 - 94 years) for orofacial cancer which almost similar with this report (19). Although the cut point for young is not universal this study found that 33.2% of cases were younger or equal to 40 year. In many literatures, incidence of orofacial cancer has been noted in younger age groups (20-22). In contrast to this finding, the research done in Chinese the median onset age is 57 years old, 51 to 70 years old is a high risk age group (23).

In this study, we have seen male predominance (57.1%), with male to female ratio of (1.48:1).This finding is almost similar with study done in Nigeria by Adesina at el showed males were (65.1%), the predominant sex affected with orofacial cancer with a 1.81:1 male to female ratio (19,24). Larizade et al. in 2014 reported that most patients (73%) were male and the overall male to female ratio was 2.74:1 (25). It seems that not only the higher rate of smoking and alcohol consumption is a vital issue but also sex hormone differences may be the reason for male predilection. More outdoor activity of males make them exposed to habits khat chewing.

Squamous cell carcinoma (SCC) was the most common type of oral and maxillofacial malignant cancer which was accounted for (50.8%) based on histologic diagnosis followed by MECA (14.2%). This result is in line with Razavi et al.’s study conducted in Iran, with the majority of SCC (60%) followed by MEC (8%) (26). Furthermore, in a UAE study (2014), the most prevalent malignant lesion was OSCC followed by MEC (27).We found, the most common malignancy of salivary glands was mucoepidermoid carcinoma (MEC) with (14.2%) followed by adenoid cystic carcinoma with (6 %). This study is comparable with the study done in west Iran, in which mucoepidermoid carcinoma is the most common salivary gland malignancies (28).

Regarding anatomic distribution, we found that oral cavity was the most common affected by oral and maxillofacial cancer, 52.6% (n=92), followed by parotid gland 14.3% (n=25) and maxillary sinus 13.7% (n=24). According to oral cavity sub site: tongue was the commonest (20%) site affected followed by lip 16 (9.1%) and buccal mucosa 14 (8%). Azimi et al, revealed lip as the most intra oral cancer (22%), followed by tongue (15%) (24).This finding is in contrast with current study. Daramola et al and Gbotolorun et al both reported the tongue as the most prevalent sub site (29.30). This finding is similar to current finding regrading to the most common anatomic sub site involvement, but still the percentage is much lower in our study subjects.

The etiology of orofacial cancer is multifactorial. In our current study, majority (68%) of patients had no knwon risk factor. Smoking in (10.3%), immune compromising disease in (14.7%), khat chewing in (6.3%) and alcoho in (1.7%) were idintified as risk factors in the study participants. Immune comporomiosing disease accounted for the majority of known risk factors in this study. Osman et al reported 49.8% tomback dipping and smoking were the major risk factors in sudan which is in contrast with this study (31). Saman Warnakulasuriya et al, on the study of Global epidemiology of oral and or pharyngeal cancer, reported smoking and alcohol has (80%) of association with cancer (32). Freidrich et al (German) reported (66.9%) of orofacial cancer case were smoker ^(33).^ These literature review support the risk factors for orofacial cancer varies globally in different regions of the world; with age, sex, habits, culture, socioeconomic and country of origin.

Most of the cases with patients with orofacial cancer (66.9%) were presented with stage IV and (17.3%) stage III. Aladelusi et al reported (65.4%) patient were diagnosed with stage IV and (34.6 %) in stage III which is similar to this study (34). Ibikunle, et al also reported that majority of patients presented with stage IV and stage III cancer (35). Friedrich et al reported 30% of stage IV cases which is lower than the current finding (36). Scott et al. compared the incidence of early-stage and advanced-stage cancer in a United States population, and reported that advanced-stage cancer was more frequent in the non-white population. This might be race and genetics are the contributing factors that play paramount role for disease progress.

According to clinical staging and primary tumour size of orofacial cancer, T4 stage was observed in the majority (53.7%) patients, followed by T3 (18.9%), T2 (21.1%), and T1 (2.3%).This clinical staging is comparable with the study done in Lahore, Pakistan, in which T4 stage was accounted for (57%) and only (2.5%) are T1 stage (37). In our current stud, 15 cases (8.6%) had distant metastasis mainly to lung. Ariyoshi et al found distant metastasis seen only in (1%) patients with orofacial cancer in Japan which is lower than this finding (38).This might be due to late presentation of patients; lack of awareness and limited service in developing country.

Surgery was a main means of treatment modality in 73(41.7%), followed by chemotherapy 43(19.4%), while 61 (34.9%) were not treated yet which is similar to the study done in Sudan (39). Delayed treatment for orofacial cancer in developing African countries including Ethiopia may be due to the limited oncologic service in government institutions and long waiting time.

To ensure quick response, early diagnosis of patients is very important. Primary health care professionals should have to have knowledge of early sign and symptoms of oral cancer in order to facilitate diagnosis and treatment before the disease become advanced. However, approximately more than half (51.14%) our study subjects were waited more than 1 year before consulting a health care professional about signs of oral cancer and only 17.71 % of patients came with less 6 months. A literature review of patients who had been referred with potentially malignant oral symptoms about 30% of patients presented to health facility after waiting three months (40). Delayed presentation to health professionals and health care facility may be influenced by lack of awareness in seeking help for orofacial cancer signs and symptoms, barriers to seeking help such as problems with access and social circumstances.

## Conclusion

According to this study majority of the cases were diagnosed with advanced stage of disease. The majority of orofacial cancer was carcinoma. Even though the cause of orofacial cancer is multifactorial, smoking, immune compromising disease, khat chewing and alcohol were some of the identified risk factor in this study. The majority of the study subjects were in stage IV AJCC with N2 where the highest lymph node metastasis. Few patients had distance metastasis to mainly lung while liver, intra-cranial and cervical vertebral metastasis were also reported. Surgery was the main means of treatment modality given to orofacial cancer patients.

Orofacial cancer is a deadly but forgotten disease in Ethiopia. The main way to address delayed presentation is through awareness creation. Early surgery is the most effective and main treatment modality for orofacial cancer. We recommend the responsible body should work in collaboration with treating physician and community leaders to increase awareness on burden of orofacial cancer. Hence capacity building of health facilities and professionals at all levels in early diagnosis and timely referral as well as improving access to oral and maxillofacial service are recommended.

## Data Availability

All relevant data are within the manuscript and its Supporting Information files

## Disclosure

### Conflict of interest

The author reports no conflicts of interest in this work.

### Author contribution

***Chala Ararsa (DMD, Oral and Maxillofacial surgeon):*** This author helped on substantial intellectual contributions to conception, design, and acquisition of data, analysis, and interpretation of data as well as on preparing the draft this study.

***Demerew Dejene (DMD, Oral and Maxillofacial surgeon):*** have made substantial contributions to conception, design, analysis and interpretation of data and participated in the critical review and editing of the manuscript drafts for scientific merit and depth.

***Gelana Garoma (DMD, Oral and Maxillofacial surgeon):*** have made substantial contributions to conception, design, analysis and interpretation of data and as well as on preparing the manuscript to this study

## Acknowledgements

The Authors would like to thank the staff and residents of Addis Ababa University, oral and maxillofacial surgery and oncology department for their support and contribution during data collection.

## Notes

### Competing Interest Statement

The authors have declared no competing interest.

### Funding Statement

The author(s) received no specific funding for this work.

### Author Declarations

Permission of ethical issue and ethical clearance was obtained from research and ethics committee of department of Dentistry, College health science, AAU before the study. This research was conducted in full accordance with the World Medical Association Declaration of Helsinki. We confirmed that patients' information remained confidential and data was anonymized and de‐identified to analysis. Patient name was not recorded and patient information kept confidential.

